# Comprehensive Angiographic Evaluation of Graft Quality After Endoscopic Vein Harvesting in Coronary Artery Bypass Grafting

**DOI:** 10.1101/2025.11.11.25340024

**Authors:** Ken Nakamura, Kentaro Akabane, Shusuke Arai, Ryota Katsura, Miku Konaka, Jun Hayashi, Eiichi Ohba, Cholsu Kim, Hideaki Uchino, Takao Shimanuki, Tetsuro Uchida

**Affiliations:** Division of Cardiovascular Surgery, Nihonkai General Hospital, Sakata, Japan; Second Department of Surgery, Yamagata University Faculty of Medicine, Yamagata, Japan

## Abstract

**Background:** The saphenous vein graft (SVG) remains a mainstay conduit for coronary artery bypass grafting (CABG) due to its accessibility and length. Although the no-touch technique may improve long-term patency, wound complications are a continuing concern. Since 2011, our institution has adopted endoscopic vein harvesting (EVH) as the standard approach. This study provides angiographic insights into graft quality and patency after EVH compared with open vein harvesting (OVH), with additional assessment of mid-term clinical outcomes.

**Methods:** Among 471 patients who underwent CABG between 2005 and 2017, 307 were included in this study. Patients were divided into the EVH group (Group A, n = 134) and the OVH group (Group B, n = 173). Postoperative coronary angiography was used to evaluate SVG graft patency, anastomotic integrity, and graft body stenosis. Clinical outcomes including major adverse cardiac and cerebrovascular events (MACCE) and wound complications were also compared.

**Results:** Angiographic assessment demonstrated comparable SVG patency between the EVH and OVH groups (93% vs. 94%), with similar rates of anastomotic stenosis (2.2% vs. 2.3%) and severe graft stenosis (≥90%; 1.5% vs. 1.2%). No significant differences were observed in 30-day mortality (1.5% vs. 3.5%), in-hospital mortality (1.5% vs. 2.1%), or postoperative stroke. Wound-related complications were rare, including wound dehiscence (1.5% vs. 2.3%) and infection (0.7% vs. 1.2%). MACCE-free survival rates at 1, 3, and 5 years were 97%, 94%, and 91% in the EVH group versus 92%, 86%, and 76% in the OVH group, respectively (P = 0.070), showing a favorable trend in the EVH group.

**Conclusion:** Detailed angiographic evaluation revealed that EVH did not compromise graft quality or patency compared with conventional OVH. The incidence of wound complications was very low, and mid-term clinical outcomes were favorable. These findings suggest that EVH is a safe and reliable harvesting technique, providing high-quality grafts with excellent angiographic integrity. Individualized selection of harvesting strategy remains important for optimizing surgical outcomes.

## Introduction

The great saphenous vein (SVG) remains the most widely used second conduit in coronary artery bypass grafting (CABG) after the internal mammary artery (IMA), owing to its adequate length and relatively low risk of vessel injury during harvesting[1, 2]. While historically associated with inferior long-term patency compared to arterial grafts, recent studies have reported favorable long-term outcomes with SVGs[3]. The no-touch harvesting technique, which avoids direct manipulation of the vein, has been proposed to improve graft durability and patency[4, 5] ; however, recent randomized controlled trials, such as the SWEDEGRAFT trial, have shown no significant difference in vein graft failure between no-touch and conventional harvesting methods, while highlighting a higher incidence of wound complications associated with the no-touch approach[6]. Endoscopic vein harvesting (EVH) has emerged as a widely adopted technique, particularly in the United States and Europe, offering significant advantages in terms of reduced wound morbidity and faster recovery[7, 8]. Unlike the no-touch method, EVH provides a minimally invasive alternative that prioritizes patient recovery and wound care. Previous studies have primarily evaluated EVH outcomes based on clinical events or overall graft patency rates, without detailed angiographic assessment of specific lesion sites. Consequently, the relationship between the harvesting technique and localized graft abnormalities—such as anastomotic narrowing, graft body stenosis, or distal runoff compromise—remains insufficiently understood.

In this study, we aimed to provide angiographic insights into graft quality and patency following EVH, directly comparing it with conventional open vein harvesting (OVH). By integrating detailed postoperative coronary angiography (CAG) findings with mid-term clinical outcomes, we sought to clarify whether EVH compromises graft integrity or durability. This comprehensive imaging-based evaluation offers a novel perspective on the safety and effectiveness of EVH in contemporary CABG practice.

## Patients and Methods

This was a retrospective observational study conducted at two centers, Nihonkai General Hospital. A total of 471 consecutive patients who underwent isolated or combined CABG between December 2005 and December 2017 were included. The study protocol was approved by the Institutional Review Board of Nihonkai General Hospital (Approval No. 007-4-12). Owing to the retrospective study design, the requirement for additional written informed consent was waived, although all patients had provided appropriate informed consent for treatment and data use at the time of care. The research was performed in compliance with the principles of the Declaration of Helsinki.

This study aimed to compare graft patency and stenosis rates between SVGs harvested using EVH and those obtained via conventional open harvesting techniques. In addition, we assessed long-term graft patency and the incidence of MACCE.

Eligible patients included those who underwent CABG with at least one SVG harvested using the EVH technique. Patients who required intraoperative conversion to open vein harvesting were excluded from the analysis. Postoperative graft assessment was conducted through qualitative evaluation using coronary angiography (CAG). Patients presenting with lower-extremity varicose veins were excluded from consideration for SVG harvesting. To evaluate the suitability of the vein, preoperative CT vein mapping was routinely performed. EVH was conducted simultaneously with the harvesting of the left internal mammary artery by an assistant surgeon, utilizing a standardized technique (VirtuoSaph system; Terumo Cardiovascular, Ann Arbor, MI). The EVH procedure—comprising skin incision, blunt tissue dissection, branch ligation, and graft preparation—was performed in accordance with established protocols previously reported in the literature[9].

The primary endpoint of this study was the incidence of early graft-related complications, such as occlusion, stenosis, or wound infection. Secondary endpoints included long-term graft patency and MACCE, defined as a composite of death, myocardial infarction, stroke, or any revascularization procedure, including coronary or non-coronary interventions (e.g., peripheral or carotid artery revascularization). Postoperative atrial fibrillation was diagnosed based on documented episodes lasting more than 30 seconds during hospitalization. Neurological events were included only when supported by findings on CT or MRI and confirmed by a neurosurgical consultation; cases suggestive of TIA without imaging evidence were excluded.

Preoperative optimization involved the management of comorbidities such as dental infections, uncontrolled diabetes mellitus, and carotid artery stenosis. All patients participated in a structured perioperative rehabilitation program under the supervision of physical therapists.

The choice between on-pump and off-pump CABG (OPCAB) was made during a multidisciplinary preoperative conference, taking into account the patient’s anatomical and clinical characteristics. On-pump CABG was preferred in cases with ventricular enlargement, reduced cardiac function, or anatomically complex targets. OPCAB was selected when complete revascularization appeared feasible on a beating heart. Conversion to cardiopulmonary bypass (CPB) was initiated intraoperatively in response to hemodynamic compromise, including ventricular arrhythmias, hypotension (systolic blood pressure ≤80 mmHg), or cardiac arrest.

During OPCAB, cardiac exposure and stabilization were achieved using posterior pericardial stay sutures, gauze packing, a tissue stabilizer (Octopus; Medtronic, Minneapolis, MN), positional adjustments, and adjuncts such as CO₂ insufflation and saline misting as needed. In on-pump CABG, the same grafting strategy was followed, with beating-heart techniques employed whenever feasible. Prophylactic intra-aortic balloon pump (IABP) support was administered preoperatively to selected high-risk patients, in accordance with indications previously reported by our institution[10].

In all cases, the left internal mammary artery was anastomosed to the left anterior descending artery. Additional revascularization of the circumflex and right coronary territories was performed using radial artery grafts or saphenous vein grafts (SVGs). A no-touch aortic technique employing bilateral internal mammary arteries was utilized in patients with suspected ascending aortic calcification or sclerosis based on imaging or intraoperative palpation. Graft quality was routinely assessed intraoperatively using a transit-time flow measurement device (Butterfly Flowmeter; Medistim, Oslo, Norway).

## Statistical analysis

Continuous variables were presented as either mean ± standard deviation or median with interquartile range, depending on data distribution. Categorical variables were summarized as counts and percentages. For comparisons of continuous variables, either the independent Student’s t-test or the Mann–Whitney U test was applied, as appropriate. Categorical data were evaluated using the chi-square test or Fisher’s exact test, as indicated.

Kaplan–Meier survival analysis was used to estimate MACCE-free survival and freedom from SVG-related events in both groups, with comparisons made using the log-rank test. All statistical analyses were performed using JMP software, version 18.2.0 (SAS Institute Japan, Tokyo, Japan).

## Results

A total of 471 consecutive patients were included in this study. Among them, 134 underwent EVH and 173 underwent OVH, as shown in Figure 1. Preoperative clinical characteristics are summarized in Table 1. There were no significant differences between the two groups in age, sex, BMI, or comorbidities such as hypertension and diabetes mellitus. However, the incidence of prior myocardial infarction was significantly lower in the EVH group compared to the OVH group (47% vs. 62%, p < 0.05). The mean follow-up duration was 21 ± 18 months in the EVH group and 47 ± 45 months in the OVH group (p < 0.0001), which likely reflects the temporal introduction of EVH beginning in 2011 at our institution. Aside from this time-related difference, baseline characteristics were largely comparable between the groups. EuroSCORE II was 2.5 ± 3.1 vs. 2.5 ± 3.0 (p = 0.982), and LVEF was 53 ± 15% vs. 54 ± 16% (p = 0.763), respectively (Table1).

**Figure 1.**
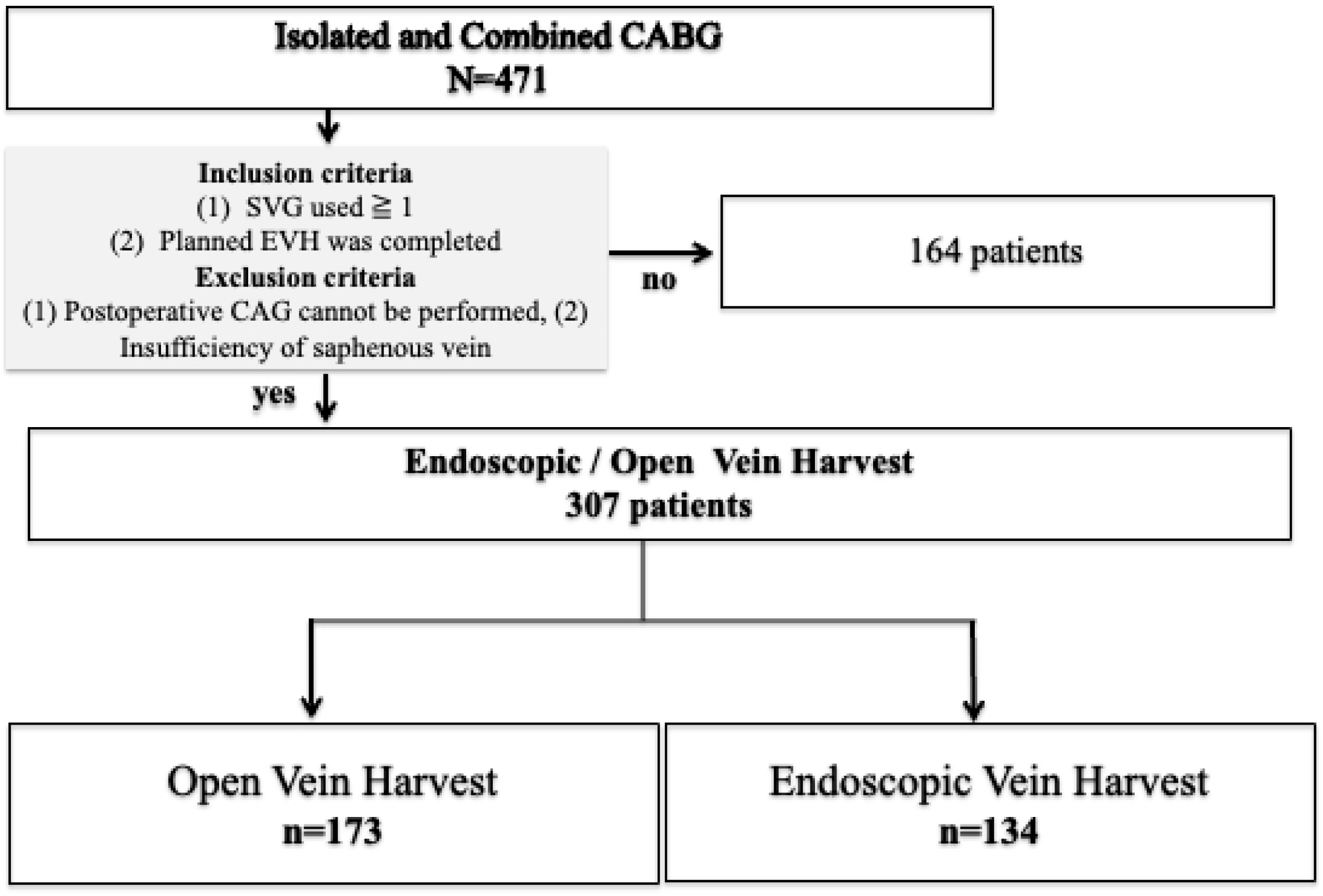
Patient Selection and Grouping Based on Vein Harvesting Technique. Among 471 patients who underwent isolated or combined coronary artery bypass grafting (CABG), a total of 307 patients met the inclusion criteria—use of ≥1 saphenous vein graft (SVG) and successful completion of planned endoscopic vein harvesting (EVH)—and did not meet the exclusion criteria—postoperative coronary angiography (CAG) not feasible or insufficient saphenous vein quality. These 307 patients were categorized into two groups based on the vein harvesting technique: 134 patients in the EVH group and 173 patients in the open vein harvesting (OVH) group. Comparative analyses were conducted between these two cohorts. CABG: coronary-artery bypass grafting, SVG: saphenous-vein graft, EVH: endoscopic vein harvesting, CAG: coronary angiography, OVH: open vein harvesting

**Table 1.**
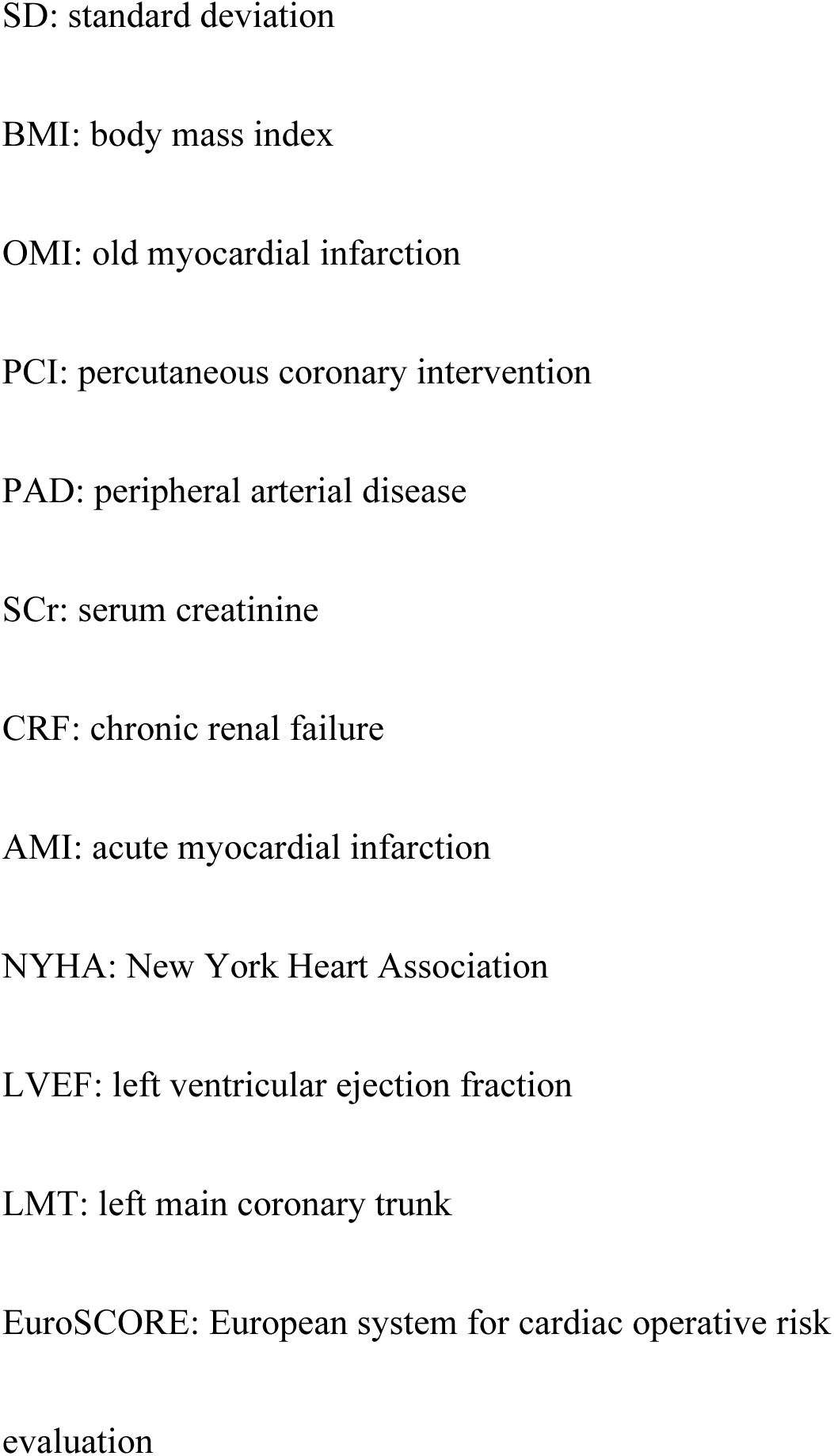
Baseline patient characteristics (preoperative data)

The number of distal anastomoses was significantly lower in the EVH group compared to the OVH group (2.7 ± 0.9 vs. 3.0 ± 0.9, p < 0.005), and fewer SVGs were used in the EVH group (1.3 ± 0.6 vs. 1.5 ± 0.7, p < 0.05). The use of LIMA was significantly more frequent in the EVH group (126/134 [93%] vs. 147/173 [85%], p < 0.05), while the use of other types of grafts did not differ significantly between the two groups.

Early postoperative SVG occlusion occurred in 9 cases (6.7%) in the EVH group and 11 cases (6.4%) in the OVH group, with no significant difference (p = 0.551). Similarly, SVG stenosis of less than 50% was observed in 1 case (0.7%) in the EVH group and none in the OVH group (p = 0.440). SVG stenosis of ≥90% was comparable between groups (2/134 [1.5%] vs. 2/173 [1.2%], p = 0.593), while stenosis of 50–90% tended to be more frequent in the EVH group, though this did not reach statistical significance (5/134 [3.7%] vs. 1/173 [0.6%], p = 0.090).

Stenosis at the SVG anastomosis site was identified in 3 cases (2.2%) in the EVH group and 4 cases (2.3%) in the OVH group (p = 0.665). Re-intervention for SVG stenosis was performed in 2 cases (1.5%) in the EVH group and 3 cases (1.7%) in the OVH group, with no significant difference (p = 0.730). Wound dehiscence was reported in 2 patients (1.5%) in the EVH group (p = 0.698), and only 1 of these cases (0.7%) was associated with wound infection (Table 2).

**Table 2.**
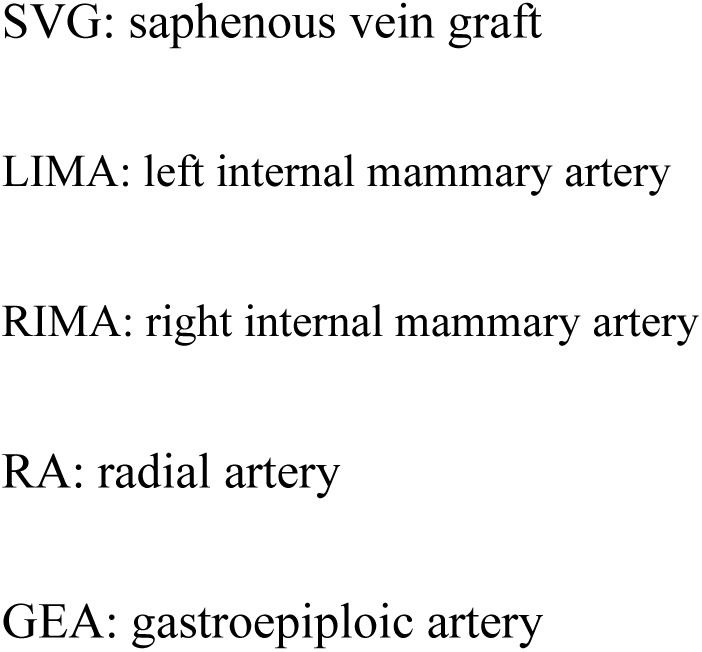
Assessment of bypass graft anastomosis and wound complication.

The EVH procedure employed in this study utilized the VirtuoSaph system (Terumo, Ann Arbor, MI), which consists of two separate components designed to independently perform vein dissection and branch ligation (Figure 2a). The skin incision required for this system is minimal and cosmetically favorable, as shown in Figure 2b, and patients rarely reported postoperative wound pain or discomfort at the harvest site.

**Figure 2.**
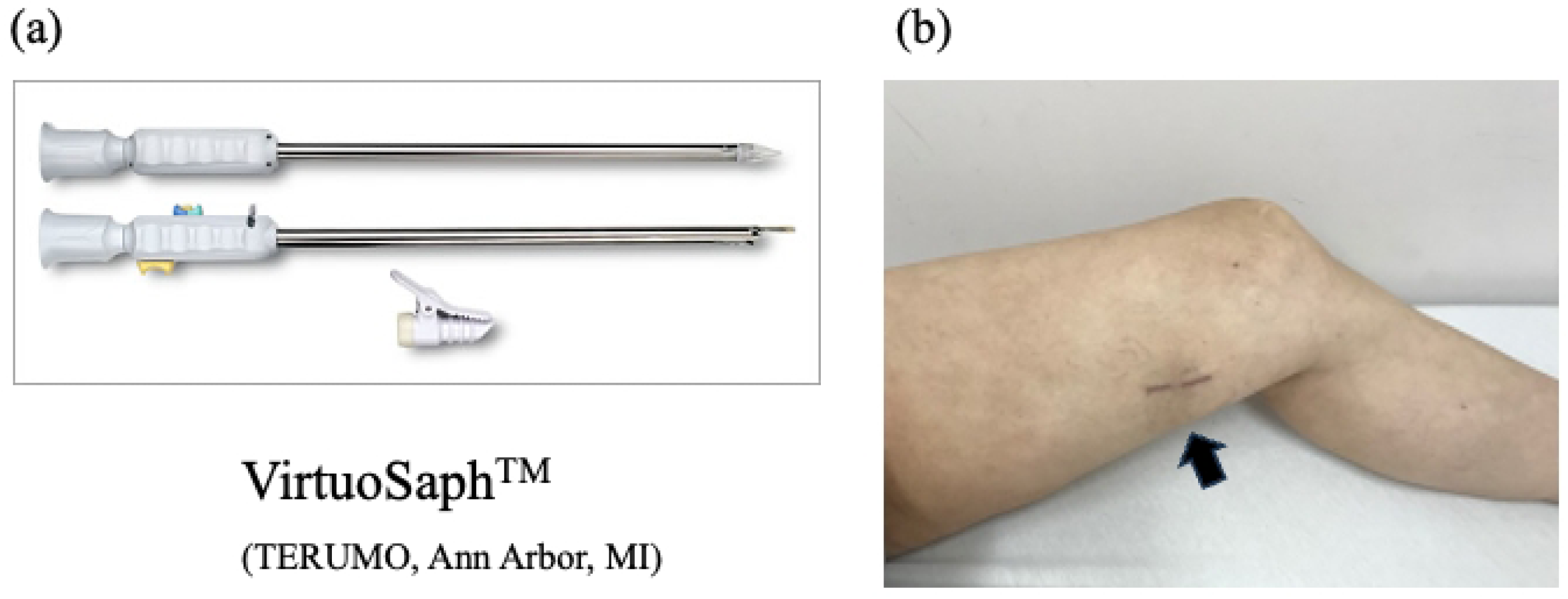
Endoscopic Vein Harvesting Using the VirtuoSaph System and Postoperative Wound Appearance. (a) The VirtuoSaph system consists of two interchangeable components used sequentially to perform skin incision, blunt tissue dissection, branch ligation, and graft preparation. (b) Postoperative view of the distal left thigh incision site. Patients rarely report pain during the postoperative course.

Intraoperative and postoperative outcomes are summarized in Table 3. No significant differences were observed between the two groups regarding the incidence of combined procedures with CABG, conversion to on-pump CABG, the need for red blood cell transfusion, peak postoperative creatinine levels, occurrence of postoperative atrial fibrillation, mediastinitis, or neurological events. Operative time was significantly shorter in the EVH group than in the OVH group (262 ± 67 min vs. 318 ± 90 min, p < 0.0001). The rate of OPCAB was also significantly higher in the EVH group (61/134 [46%] vs. 55/173 [33%], p < 0.05). CPB time tended to be shorter in the EVH group (137 ± 45 min vs. 150 ± 57 min, p = 0.065), though the difference was not statistically significant. There were no significant differences in the duration of mechanical ventilation (1.3 ± 1.6 days vs. 1.5 ± 1.9 days, p = 0.270) or ICU stay (4.7 ± 3.2 days vs. 5.2 ± 5.7 days, p = 0.427). However, postoperative hospital stay was significantly shorter in the EVH group (22 ± 11 days vs. 26 ± 21 days, p = 0.019).

**Table 3.**
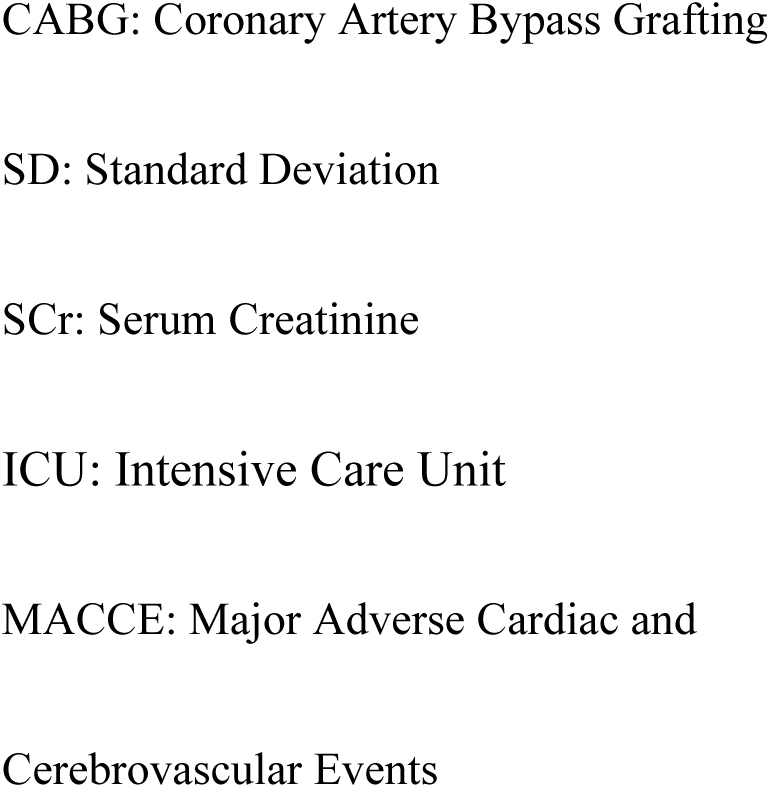
Clinical Outcomes and Complications According to Vein Harvesting Technique.

The 30-day mortality was 1.5% in the EVH group and 3.5% in the OVH group (p = 0.473), and in-hospital mortality was 1.5% vs. 4.7% (p = 0.194). One-year postoperative MACCE occurred significantly less frequently in the EVH group (4% vs. 21%, p < 0.001) (**table 3**).

Cardiac death occurred in 1.5% of patients in the EVH group and 4.7% in the OVH group (p = 0.194). No fatal events were reported after hospital discharge in either group. The SVG-related event-free survival at 1, 3, and 5 years was identical between the groups: 93%/93%/93% in the EVH group and 93%/93%/93% in the OVH group (p = 0.919), with no significant differences observed (**Figure 3**). In the Kaplan-Meier analysis for MACCE-free survival, the event-free rates at 1, 3, and 5 years were 97%, 94%, and 91% in the EVH group, and 92%, 86%, and 76% in the OVH group, respectively (p = 0.070), showing a favorable trend in the EVH group (**Figure 4**).

**Figure 3.**
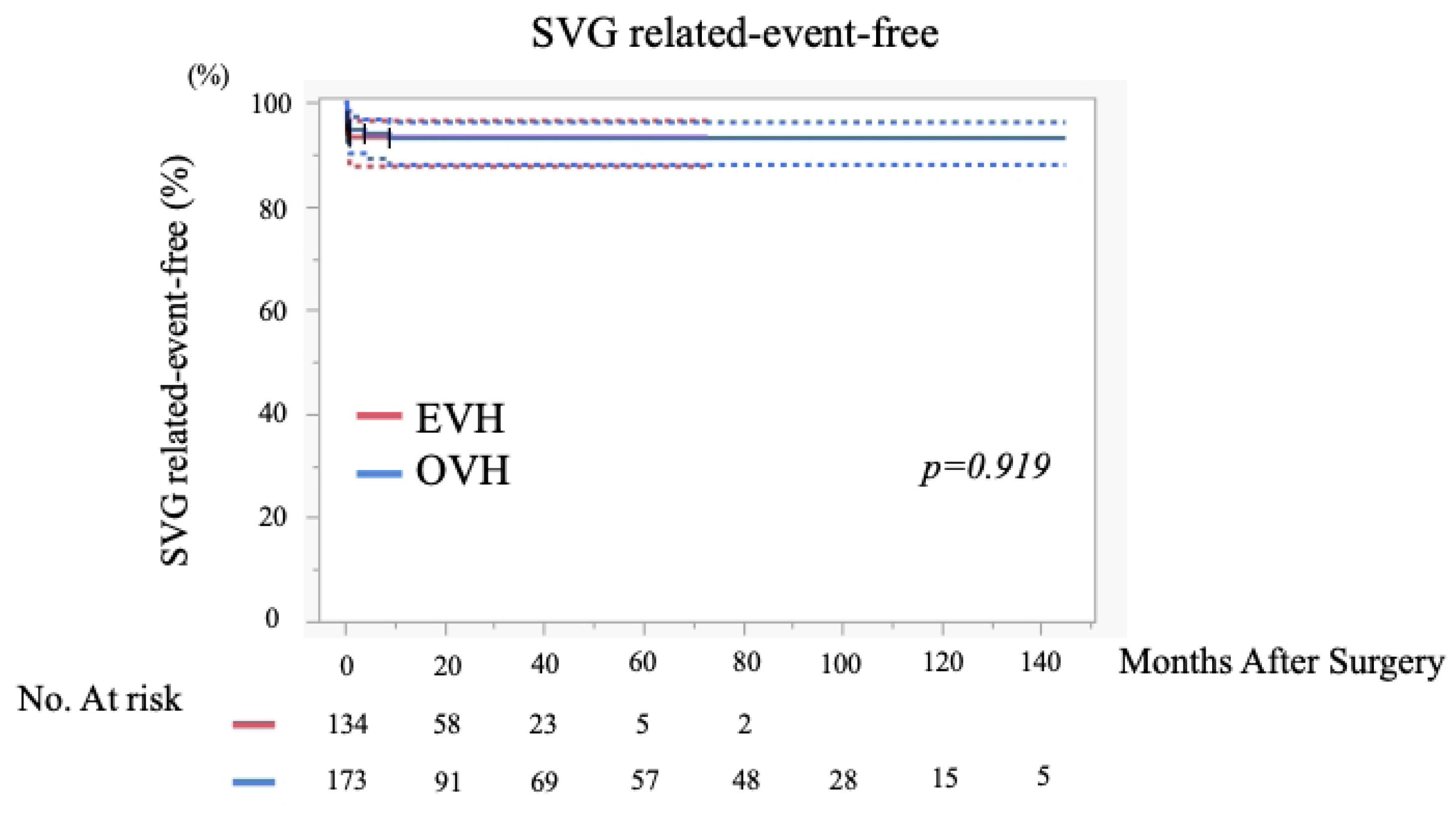
Kaplan–Meier Curves for SVG-Related Event-Free Survival in 307 CABG Patients. Kaplan–Meier analysis of saphenous vein graft (SVG)-related event-free survival in 307 patients who underwent isolated or combined coronary artery bypass grafting (CABG) at our institution. Patients were divided into two groups: 134 who underwent endoscopic vein harvesting (EVH) and 173 who underwent open vein harvesting (OVH). Follow-up was continued for up to 140 months, and differences between the groups were evaluated using the log-rank test. SVG: saphenous-vein graft, CABG: coronary-artery bypass grafting, EVH: endoscopic vein harvesting, OVH: open vein harvesting

## Discussion

SVG plays an essential role as the second conduit in CABG following IMA, and its quality may directly influence long-term survival and the incidence of major cardiovascular events. In addition to conventional open vein harvesting, techniques such as the no-touch method—designed to preserve the perivascular tissue—have been associated with improved long-term patency[4, 11]. Meanwhile, EVH has gained attention due to its advantages in reducing wound-related complications, such as infection and pain. Recent studies suggest that the method of SVG harvesting may significantly impact both graft patency and clinical outcomes after CABG[12, 13].

The present study yielded several important insights: 1) EVH did not demonstrate any inferiority compared to OVH in terms of graft quality, as evidenced by comparable rates of SVG stenosis and occlusion on follow-up imaging. 2) The incidence of wound-related complications associated with EVH was low, occurring in less than 2%, and no cases of refractory or deep wound infections were observed. 3) There was no indication that EVH had a negative impact on early or midterm postoperative outcomes in patients undergoing CABG.

These findings are consistent with previous reports suggesting that EVH can maintain graft quality while significantly reducing wound-related complications. Studies such as PREVENT IV and others have previously raised concerns regarding EVH and long-term graft patency [14, 15], but more recent analyses and meta-analyses have reported that when EVH is performed using a standardized technique, clinical outcomes and graft patency are comparable to those of OVH[16, 17]. Our results further support the notion that EVH offers a safe and effective conduit harvesting strategy in contemporary CABG. A key strength of our study lies in the detailed qualitative assessment of SVGs performed in the early postoperative period using CAG. This included evaluation of graft body stenosis and anastomotic site stenosis, an approach that has been rarely reported in the existing literature. In a study by J. Ran et al., the incidence of early SVG occlusion was reported as 11.6% in the EVH group and 9.8% in the OVH group, with no significant difference between the two techniques. Similarly, in our study, the rates were 6.7% for EVH and 6.4% for OVH, which are comparable and consistent with these previous findings[18]. Another report indicated that graft patency rates between EVH and OVH remained similar up to 6 months postoperatively, with a decline in EVH graft patency observed at 12 months. However, that study did not include a detailed anatomical or morphological evaluation of the grafts, and the reason for the divergence in patency after 12 months remains unclear[19].

While the superiority of EVH over OVH in terms of postoperative wound pain, wound healing, and infection has become widely accepted and is no longer a matter of debate, the underlying mechanisms continue to be explored. Notably, lymphoscintigraphic data have suggested that improved lymphatic drainage in EVH may account for its favorable outcomes, offering a micro-level explanation for the reduced incidence of wound-related complications[20].

In parallel, numerous studies have focused on improving long-term outcomes through the use of no-touch saphenous vein grafts (NT-SVG). However, EVH and NT-SVG harvesting techniques offer distinctly different advantages and limitations. Given their differing profiles, the selection of an appropriate harvesting method should be tailored to the individual patient’s clinical characteristics, which may enhance overall surgical outcomes[6, 12].

Several limitations of this study should be acknowledged. First, the sample size was relatively small, which may limit the statistical power of the findings. Second, patient selection was not randomized; candidates for EVH were chosen at the discretion of the operating surgeons, potentially introducing selection bias. Third, due to the non-randomized study design, unmeasured confounding factors and detection bias may have influenced the results. Fourth, long-term postoperative coronary angiography was not performed in all patients, which limited the ability to conduct detailed qualitative assessments of the grafts during the late phase. Because follow-up angiography was performed primarily in patients with suspected coronary events, the actual rate of graft-related complications may have been underestimated.

Furthermore, future evaluations of the program should include cost-effectiveness analyses regarding hospitalization and procedural costs, to comprehensively assess the benefits of EVH. Lastly, the generalizability of our findings to other institutions or international settings remains uncertain, and external validation is warranted.

## Conclusion

EVH for SVG harvesting did not negatively affect graft quality or patient survival. Wound-related complications such as poor healing and infection occurred at a very low rate. These results suggest that EVH is a safe and reliable technique. Flexible selection of harvesting methods based on each patient’s background and comorbidities appears to be essential for optimizing surgical outcomes.

## Acknowledgments

This study did not receive any specific support.

## Author Contributions

Conceptualization: Ken Nakamura, Hideaki Uchino, Takao Shimanuki, Tetsuro Uchida

Data curation: Ken Nakamura, Kentaro Akabane, Shusuke Arai, Ryota Katsura, Miku Konaka, Jun Hayashi, Eiichi Ohba

Formal analysis: Ken Nakamura

Investigation: Ken Nakamura, Tetsuro Uchida

Methodology: Ken Nakamura, Cholsu Kim, Hideaki Uchino, Tetsuro Uchida

Project administration: Ken Nakamura, Shusuke Arai

Resources: Ken Nakamura

Supervision: Takao Shimanuki, Tetsuro Uchida

Writing –original draft: Ken Nakamura

Writing-review and editing: Ken Nakamura, Hideaki Uchino, Tetsuro Uchida

## Data Availability Statement

Data is provided within the supplementary information file.

## Ethics, Consent to Participate, and Consent to Publish declarations

This study was reviewed and approved by the Institutional Review Board of Nihonkai General Hospital (Approval No. 007-4-12). Appropriate informed consent for treatment and data use was obtained from all participants. However, the requirement for additional written informed consent to participate in this retrospective study was waived by the committee. The research was conducted in accordance with the principles outlined in the Declaration of Helsinki.

## Consent for publication

Not Applicable

---

**Figure 3.** Kaplan–Meier Curves for MACCE-Free Survival in 307 CABG Patients.

Kaplan–Meier curves showing major adverse cardiac and cerebrovascular event (MACCE)-free survival in 307 patients who underwent isolated or combined coronary artery bypass grafting (CABG) at our institution. All saphenous vein grafts (SVGs) were harvested using either endoscopic vein harvesting (EVH; n = 134) or open vein harvesting (OVH; n = 173). Comparison between the two groups was performed using the log-rank test.

MACCE: major adverse cardiac and cerebrovascular events, CABG: coronary-artery bypass grafting, SVG: saphenous-vein graft, EVH: endoscopic vein harvesting, OVH: open vein harvesting

